# Population-based stroke surveillance using Big Data: epidemiological trends in admissions and mortality in Australia

**DOI:** 10.1101/2020.01.08.20016899

**Authors:** Melina Gattellari, Chris Goumas, Bin Jalaludin, John M. Worthington

## Abstract

**Background:** Epidemiological trends for major causes of death and disability, such as stroke, may be monitored using administrative data to guide public health initiatives and service delivery.

**Methods:** We calculated admissions rates for ischaemic stroke, intracerebral haemorrhage or subarachnoid haemorrhage between January 1, 2005 and December 31^st^, 2013 and rates of 30-day mortality and 365-day mortality in 30-day survivors to December 31^st^ 2014 for patients aged 15 years or older from New South Wales, Australia (population 7.99 million). Annual Average Percentage Change in rates was estimated using negative binomial regression.

**Results:** Of 81,703 eligible admissions, 64,047 (78.4%) were ischaemic strokes and 13,302 (16.3%) and 4,778 (5.8%) were intracerebral and subarachnoid haemorrhages, respectively. Intracerebral haemorrhage admissions significantly declined by an average of 2.2% annually (95% Confidence Interval=-3.5% to −0.9%) (p<0.001). Thirty-day mortality rates significantly declined for ischaemic stroke (Average Percentage Change −2.9%, 95% Confidence Interval=-5.2% to −1.0%) (p=0.004) and subarachnoid haemorrhage (Average Percentage Change=-2.6%, 95% Confidence Interval=-4.8% to −0.2%) (p=0.04). Mortality at 365-days amongst 30-day survivors of ischaemic stroke and intracerebral haemorrhage was stable over time and increased in 30-day survivors of subarachnoid haemorrhage (Annual Percentage Change 6.2%, 95% Confidence Interval=-0.1% to 12.8%), although the increase was not statistically significant (p=0.05).

**Conclusion:** Improved prevention may have underpinned declining intracerebral haemorrhage rates while gains in survival suggest that innovations in stroke care are being successfully translated. Longer-term mortality in patients surviving the acute period is unchanged and may be increasing for subarachnoid haemorrhage warranting investment in post-discharge care and secondary prevention.

## Introduction

Stroke is a major cause of death and disability.^1^ In well-resourced countries, the burden of stroke has increased due to an ageing population,^2^ while stroke incidence has more than doubled over the past four decades in low to middle income countries.^3^

Worldwide, there is growing impetus to dedicate resources to reduce the adverse impact of stroke on survival and quality of life.^2,4^ Anti-hypertensives and statins are proven to prevent stroke,^5^ while evidence-based innovations in acute care, including interventional neuroradiology for subarachnoid haemorrhage, improved access to comprehensive stroke unit care, thrombolysis for ischaemic stroke and, more recently, mechanical thrombectomy for large vessel occlusions have been implemented in several countries reducing the likelihood of death and/or disability.^4^

Monitoring epidemiological trends is essential to determine the impact of initiatives for stroke prevention and in-hospital management, to guide future care and inform resource allocation.^6^ Routinely collected health data may usefully address this need as in several jurisdictions these are mandated collections capturing hospitalisations for a geographically defined population and have done so continuously since their inception. “Big data” is enhanced with linkages between and across data-sets, allowing prospective individual-level analyses to measure post-discharge outcomes, including longer-term mortality.^7^

We examined trends in stroke hospitalisations and survival, using routinely collected linked hospital admissions and mortality data within the context of an upscaling and intensification of resources in Australia to prevent and manage stroke. Our analyses focussed on hospital presentations to address the burden of stroke on health services and to examine trends in patient outcomes after hospitalisation.

## Methods

Analyses were conducted as part of the Home to Outcomes (H20) study,^8-10^ a data linkage project reporting stroke epidemiology and outcomes in New South Wales (NSW), Australia’s most populous state (population ∼7.99 million in 2019).^11^ The study was approved by the NSW Population Health Services Research Ethics Committee (HREC/14/CIPHS/17). In part, the analyses reported here update those carried out for a previous data-linkage study in which we reported epidemiological trends for haemorrhagic stroke sub-types in New South Wales from 2001 to 2009. ^12,13^

As described previously,^8-10^ patients over 15 years of age were identified from the Admitted Patient Data Collection, a census of all hospitalisations in NSW, including deaths occurring within the emergency department, recording the reason for an admission (Principal diagnosis) and up to 49 secondary diagnoses. Trained coders apply the International Classification of Diseases, version 10, Australian Modification (ICD-10AM)^14^ to abstract medical records after patient discharge according to standardised criteria.

We selected cases with a principal stroke diagnosis recorded from January 1, 2005 to December 31^st^, 2013. Discharges were available until June 30, 2014, allowing at least six-months for admissions towards the end of 2013 to be coded. We identified cases of subarachnoid haemorrhage (ICD-10AM I60), intracerebral haemorrhage (ICD-10AM I61, I62.9) and ischaemic stroke (ICD-10AM I63, I64) applying recommended ICD-10 codes.^15,16^ Acute strokes recorded in secondary diagnostic positions were included where the primary diagnosis was consistent with an acute stroke event (for example, cardiac arrhythmia, hemiplegia), where iatrogenic causes were identified, or if flagged as an in-hospital event.

Separations indicating either inter- or intra-hospital transfers, a change in the type of care provided (for example, from acute to sub-acute care), death or discharge to home or other accommodation, were linked to form a continuous period of hospitalisation. As previously described,^8^ contiguous episodes were joined if separated by up to 24 hours^17^ or if recorded by the end of the next day if the preceding separation was recorded at another hospital.

Admissions were linked to death registrations obtained from the NSW Registry of Deaths, to December 31^st^, 2014, to ascertain 30- and 365-day mortality. Probabilistic and deterministic linkage was carried out by a government provider using gold-standard privacy preserving protocols.^18^ The false-positive linkage rate did not exceed 5 per 1,000 linkages.

### Case selection

We excluded patients residing outside NSW to define the geographical population-base and patients whose records included concomitant diagnoses of traumatic cerebral haemorrhage (ICD-10AM codes S04, S06, S07, S08, S17, S18, S01.7, S02.0, S02.1, S02.6, S02.7, S03.0, S09.0, S09.2, S09, S05.2, S05.7) or cerebral neoplasms (ICD-10AM C71, C70.0,C70.9,C79.3,D33.0,D33.1,D33.2,D33.3,D33.9). Cases with a principal stroke diagnosis revised after early transfer to another hospital by the end of the next day after presentation were excluded as presumed misdiagnosed cases. We also excluded cases discharged home before 48 hours consistent with recommendations to improve case specificity^19^ because such cases might represent unrevised provisional cases, stroke mimics or those with rapidly resolving symptoms after a transient ischaemic attack. As these cases may also indicate mild strokes, particularly milder ischaemic strokes, sensitivity analyses for assessing temporal trends included these early discharges.

### Statistical analyses

Stroke admission rates were directly standardised to the World population,^20^ grouped by calendar year and five-year age groups. We calculated crude and directly standardised rates of fatal strokes for cases dying within 30-days of admission and for 365-day mortality in 30-day survivors using the NSW population as the denominator.^11^ The average annual percentage change (APC) was estimated using negative binomial regression models, modelling the number of cases as the outcome, offset against the natural logarithm of the age-, sex- and year-specific population estimates, with calendar year fitted as a linear term, adjusting, where applicable, for sex and age within ten-year age groups. We tested interaction terms to determine if trends differed according to sex and age and analysed trends for each subtype and also for ischaemic stroke and intracerebral haemorrhage combined, representing strokes largely amenable to risk modification^5^ and stroke unit care.^4^ P-values less than 0.05 were deemed statistically significant.

## RESULTS

### Cohort selection and characteristics

We identified 81,703 eligible admissions (Supplementary Figure 1), of which 64,047 (78.4%) were classified as ischaemic strokes, while 13,302 (16.3%) and 4,778 (5.8%) were intracerebral and subarachnoid haemorrhages, respectively.

The median age was 78 and 77 years, respectively for ischaemic stroke and intracerebral haemorrhage patients, while patients with subarachnoid haemorrhage had a median age of 58 years. Males and females were equally represented amongst patients with intracerebral haemorrhage, while 60.9% of subarachnoid haemorrhage cases were women. Men were slightly over-represented amongst ischaemic stroke admissions (51.2%) (Table 1).

**Table 1.** Patient characteristics by stroke sub-type

|  | Ischaemic |  | ICH |  | SAH |  |
| --- | --- | --- | --- | --- | --- | --- |
|  | N | % | N | % | N | % |
| Age at diagnosis |  |  |  |  |  |  |
| Mean age | 75 | SD=13 | 73 | SD=15 | 59 | SD=17 |
| Median age | 78 | IQR=68-85 | 77 | IQR=66-84 | 58 | 66-84 |
| Sex |  |  |  |  |  |  |
| Male | 32,761 | 51.2 | 6,498 | 50.0 | 1,828 | 39.1 |
| Female | 31,286 | 48.8 | 6,488 | 50.0 | 2,842 | 60.9 |
| **Total cases** | **64,047** | **(100)** | **12,986** | **(100)** | **4,670** | **(100)** |
| Fatal stroke in 30-days | 10,415 | 16.3 | 5,011 | 38.6 | 1,198 | 25.7 |
| 30-day survivors | 53,632 | 83.7 | 7,975 | 61.4 | 3,472 | 74.3 |
| 365-day mortality in 30-survivors | 8,279 | 15.4 | 1,260 | 15.8 | 179 | 5.2 |

ICH=Intracerebral Haemorrhage
SAH=Subarachnoid Haemorrhage

The crude rate of ischaemic stroke admissions was estimated as 125 per 100,000 population and the age-standardised rate was 75 per 100,000. The respective crude and age-standardised estimates were 25 and 16 for intracerebral haemorrhage and 9 and 7 per 100,000 for subarachnoid haemorrhage (Table 2).

**Table 2:** Age standardised admission rates (ASR per 100,000) by year and stroke sub-type

|  |  | IS |  |  | ICH |  |  | SAH |  |  | IS & ICH |  |  |
| --- | --- | --- | --- | --- | --- | --- | --- | --- | --- | --- | --- | --- | --- |
| Year | Person Years | N | Crude Rate | ASR 95% CI | N | Crude Rate | ASR 95% CI | N | Crude Rate | ASR 95% CI | N | Crude Rate | ASR 95% CI |
| 2005 | 5,373,880 | 7,287 | 136 | 85 (83-87) | 1,410 | 26 | 17 (16-18) | 464 | 9 | 7 (7-8) | 8,697 | 162 | 102 (100-104) |
| 2006 | 5,423,700 | 7,171 | 132 | 82 (80-84) | 1,534 | 28 | 18 (17-19) | 503 | 9 | 8 (7-8) | 8,705 | 160 | 100 (98-102) |
| 2007 | 5,507,371 | 7,068 | 128 | 79 (77-81) | 1,347 | 24 | 16 (15-17) | 497 | 9 | 7 (7-8) | 8,415 | 153 | 95 (93-97) |
| 2008 | 5,606,333 | 7,098 | 127 | 77 (75-79) | 1,434 | 26 | 16 (15-17) | 541 | 10 | 8 (7-9) | 8,532 | 152 | 93 (91-95) |
| 2009 | 5,704,008 | 6,990 | 123 | 75 (73-76) | 1,425 | 25 | 16 (15-17) | 536 | 9 | 8 (7-8) | 8,415 | 148 | 90 (88-92) |
| 2010 | 5,783,499 | 6,777 | 117 | 70 (69-72) | 1,448 | 25 | 16 (15-17) | 496 | 8 | 7 (6-8) | 8,225 | 142 | 86 (84-88) |
| 2011 | 5,851,202 | 7,101 | 121 | 71 (69-73) | 1,446 | 25 | 15 (14-16) | 530 | 9 | 7 (7-8) | 8,547 | 146 | 86 (84-88) |
| 2012 | 5,926,116 | 7,305 | 123 | 72 (70-74) | 1,485 | 25 | 15 (15-16) | 552 | 9 | 8 (7-8) | 8,790 | 148 | 88 (86-90) |
| 2013 | 6,009,142 | 7,250 | 121 | 70 (69-72) | 1,457 | 24 | 14 (14-15) | 551 | 9 | 7 (7-8) | 8,707 | 145 | 85 (83-87) |
| All years | 51,185,251 | 64,047 | 125 | 75 (75-76) | 2,986 | 25 | 16 (16-16) | 4,670 | 9 | 7 (7-8) | 77,033 | 150 | 91 (91-92) |

IS=Ischaemic Stroke; ICH=Intracerebral Haemorrhage; SAH=Subarachnoid Haemorrhage

### Admission trends

While admission rates declined for all subtypes over the nine-year study period, statistically significant decreases were identified only for intracerebral haemorrhage (Table 3). Age-standardised rates for intracerebral haemorrhage decreased from 17 to 14 per 100,000 from 2005 to 2013, resulting in a significant average decrease of 2.2% per year (95% CI=-3.5% to −0.9%) (p<0.001). The age-standardised admission rate for ischaemic stroke decreased from 85 per 100,000 to 70 per 100,000 in 2013. However, the adjusted APC decline of 1.2% per year (95% CI=-2.4 to 0.1) did not reach significance (p=0.08). When admissions for ischaemic strokes and intracerebral haemorrhages were combined, rates significantly decreased by an average of 1.4% per year (95% CI=-2.6% to −0.2%) (p=0.02). Results from sensitivity analyses including discharges to home within 48 hours were consistent with main analyses. However, the trend in intracerebral haemorrhages and ischaemic strokes combined was marginally non-significant (p=0.09) (Table 3).

**Table 3.** Average percentage change (APC) per year in admission rates

| Sub-type | Main Analysis |  | Sensitivity Analysis<sup>a</sup> |  |
| --- | --- | --- | --- | --- |
|  | APC (95% CI) | p-value | APC (95% CI) | p-value |
| **Admission rates** |  |  |  |  |
| Ischaemic Stroke | -1.2 (-2.4 to 0.1) | 0.08 | -0.7 (-2.0 to 0.6) | 0.26 |
| Intracerebral Haemorrhage | -2.2 (-3.5 to -0.9) | <0.001 | -2.1 (-3.3 to -0.8) | 0.001 |
| Subarachnoid Haemorrhage | -0.2 (-1.4 to 1.0) | 0.73 | -0.0 (-1.2 to 1.2) | 0.96 |
| Ischaemic stroke and intracerebral haemorrhage | -1.4 (-2.6 to -0.2) | 0.02 | -1.0 (-2.2 to 0.2) | 0.09 |
| **30-day fatality rates** |  |  |  |  |
| Ischaemic Stroke | -3.1 (-5.2 to -1.0) | 0.004 | -2.9 (-5.0 to -0.8) | 0.007 |
| Intracerebral Haemorrhage | -1.9 (-3.9 to 0.1) | 0.06 | -1.8 (-3.8 to 0.2) | 0.08 |
| Subarachnoid Haemorrhage | -2.6 (-4.9 to -0.2) | 0.04 | -2.5 (-4.9 to -0.1) | 0.04 |
| Ischaemic stroke and intracerebral haemorrhage | -2.6 (-4.5 to -0.8) | 0.006 | -2.4 (-4.3 to -0.5) | 0.01 |
| **365-day fatality rates<sup>b</sup>** |  |  |  |  |
| Ischaemic Stroke | 0.3 (-2.6 to 3.4) | 0.82 | 0.3 (-2.7 to 3.3) | 0.86 |
| Intracerebral Haemorrhage | -0.2 (-3.3 to 3.0) | 0.91 | 0.0 (-3.0 to 3.2) | 0.98 |
| Subarachnoid Haemorrhage | 6.2 (-0.1 to 12.8) | 0.05 | 6.1 (-0.1 to 12.7) | 0.05 |
| IS&ICH | 0.4 (-3.3 to 4.2) | 0.84 | 0.3 (-3.3 to 4.1) | 0.86 |

<sup>a</sup>Includes cases discharged alive within 48 hours; <sup>b</sup>In 30-day survivors

For both ischaemic stroke and intracerebral haemorrhage admissions, separately and in combination, rates significantly declined for cases over 75 years of age when tested as a discrete age-group. However, trends did not significantly differ according to age and sex when interaction terms were tested (Table 3, Supplementary Table 1).

### Trends in mortality rates following hospital admission

There were 16,624 deaths within 30-days (20.3%). The median age of fatal cases was 84 years (Quartile 1=78; Quartile 3=89) for ischaemic stroke, 81 years (Quartile 1=72 and Quartile 3=86) for intracerebral haemorrhage and 73 years (Quartile 1=57 and Quartile 3=83) for subarachnoid haemorrhage (Table 1).

Almost one in five (19.5%) females and 13.2% males died within 30-days after an ischaemic stroke while 41.4% and 35.8% of intracerebral haemorrhage admissions ended in death within 30-days for females and males, respectively. Thirty-day mortality in subarachnoid haemorrhage cases was 27.4% amongst females and 22.9% amongst males.

Thirty-day mortality rates declined for all subtypes (Table 3, Figure 1). Amongst ischaemic stroke admissions, age-standardised mortality rates fell from 11.6 per 100,000 in 2005 to 8.4 per 100,000 in 2013, an average annual percentage decrease of 2.9% (95% CI=-5.2 to −1.0) (p=0.004). An average annual decline of 1.6% per year in intracerebral haemorrhage mortality (95% CI=-3.9% to 0.1%) neared statistical significance (p=0.06). The estimated average annual percentage decline in 30-day mortality after subarachnoid haemorrhage was 2.6% (95%CI=-4.9 to −0.2) (p=0.04). When ischaemic and intracerebral haemorrhage admissions were combined, mortality fell by an average of 2.4% per year (95% CI=-4.3 to −0.5), (p=0.0006). Results were unchanged when discharges within 48 hours were included.

**Figure 1:**
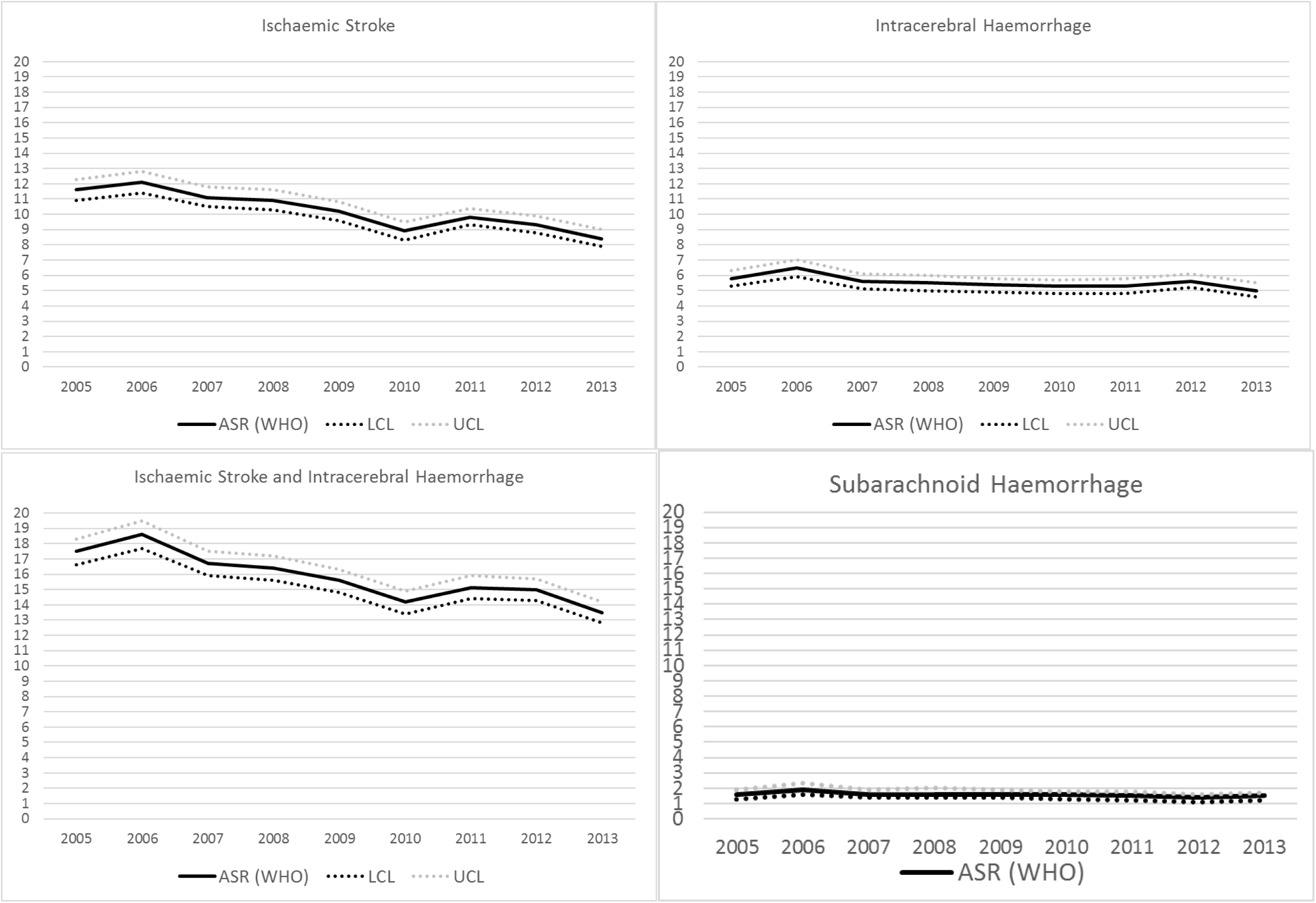
Trends in fatality rates 30-days after stroke (per 100,000) age-standardised to the World Health Organisation (WHO) World Population.

Ischaemic stroke fatality rates significantly decreased for both sexes while death after subarachnoid haemorrhage significantly declined for females only. Amongst ischaemic stroke admissions, declines were largest for the 75 to 84-year age group. Amongst subarachnoid haemorrhage admissions, patients aged 65-74 years experienced the largest average decline of 8.2% per year (95% CI=-13.8 to −2.2) (p=0.008). Interaction terms testing whether trends significantly varied according to age or sex, however, were not statistically significant for any subtype (Supplementary Table 3).

### Trends in 365-day mortality rates amongst 30-day survivors

Amongst the 65,079 patients surviving beyond 30-days, 9,718 died within 365-days (14.9%) of the stroke admission. One in eight 30-day survivors of ischaemic stroke (15.4%, n=8,279 53,632) and intracerebral haemorrhage (15.8%, n=1,260 out of 7,975) died within 365-days. Death within a year in 30-day survivors of subarachnoid haemorrhage occurred in 5.2% of cases (179 out of 3,472) (Table 1).

Mortality at 365-days amongst 30-day survivors was static over time for ischaemic strokes and intracerebral haemorrhages. There was evidence that 365-day mortality in 30-day survivors increased for subarachnoid haemorrhage (APC=6.2%, 95% CI=-0.1% to 12.8%), a marginally non-significant finding (p=0.05). There was no evidence these trends varied by age or sex. Sensitivity analyses including suspected provisional stroke cases discharged within 48 hours replicated these findings (Table 3).

## Discussion

### Main findings: Admission trends

We observed stable admission rates of subarachnoid haemorrhage, non-significant declines for ischaemic stroke and significantly reduced rates of intracerebral haemorrhage over the study period. These results suggest on-going challenges in preventing ischaemic stroke and subarachnoid haemorrhage in our community.

Increasing use of anti-hypertensives may have driven declining rates of intracerebral haemorrhage.^21^ The decline in intracerebral haemorrhage risk is reassuring given anticoagulants were more widely used in Australia towards the end of the study period^22^ although the impact of this on haemorrhagic stroke risk is yet to be researched in our setting.

Age-standardised admission rates of ischaemic stroke decreased from 85 to 70 per 100,000 over the study period, although the reduction was not significant. Larger and significant declines were observed for patients older than 75 years of age, a cohort most likely targeted for primary prevention. While these results suggest that reductions in risk are being realised, the overall non-significant trend is potentially concerning given ischaemic strokes are largely preventable. Diagnostic upstaging of transient ischaemic attack due to wider use of magnetic resonance imaging over time may have mitigated against observing reductions in risk. Moreover, evidence-based protocols for thrombolysis permit treatment of clinical signs in the absence of positive imaging.^23^ Over time, an increasing number of patients with stroke mimics or transient ischaemic attacks may have been identified and treated as stroke cases.

Rates of subarachnoid haemorrhage admissions were unchanged. The main risk factors, namely female sex and advancing age, are non-modifiable. Further, there is no established means to prevent the development of aneurysms and screening to identify and secure unruptured aneurysms in high risk groups, such as those with a family history, is not established. Therefore, there is little reason to expect changing community risk of subarachnoid haemorrhage. Our findings did not replicate Finnish^24^ and Irish^25^ studies, both utilising administrative data reporting declining subarachnoid haemorrhage rates in tandem with decreasing rates of smoking over 14^24^ and 18^25^ years. In NSW, the proportion of adults smoking daily has decreased from 16.0% to 12.0% between 2005 and 2013^26^ with no observable impact on subarachnoid haemorrhage admissions, possibly because declines in relatively low smoking rates may have been insufficient to affect a comparatively low risk of subarachnoid haemorrhage. We note population attributable risk percentages for hypertension, smoking and alcohol intake exceed or approach 20%^27^ arguing for concerted initiatives to prevent subarachnoid haemorrhage.

### Main findings: Mortality rates

Evidence supporting declining mortality was more convincing. Two plausible reasons for genuine improvements in survival may underpin these results. First, increasing uptake of preventative treatments may have resulted in milder, less fatal and disabling strokes.^28^ Second, during the study period, NSW saw expansion of stroke unit care and thrombolysis,^29^ and the implementation of a state-wide network offering around the clock neurosurgical care, including endovascular coiling. We previously demonstrated declining mortality after subarachnoid haemorrhage in concert with an increasing uptake of endovascular coiling as reported in a previous data-linkage study.^13^

One-year mortality in 30-day survivors of ischaemic stroke and intracerebral haemorrhage was stable suggesting the wider use of life-preserving treatment did not postpone death beyond the acute period. Major advances in stroke care and outcomes in recent years have concentrated on hyper-acute and acute care while the post-acute phase has received less attention from researchers and health care managers when re-designing stroke services. A key question for future research is to determine to what extent complications of stroke and the risk of recurrent stroke contribute to longer-term mortality.

There was a near significant increase in 365-day mortality in 30-day survivors after a subarachnoid haemorrhage. This may represent a “cost of treatment” for pursuing life-sustaining interventions in relatively young patients with likely high premorbid functioning where the evidence-base for early intervention is conclusive. More aggressive treatment may be offered by clinicians and accepted or requested by patients and/or their families even when there is limited hope for substantive recovery. Such “cost” may be difficult to avoid as accurate longer-term prognostication is challenging in the acute critical phase where treatment must be promptly administered before the full extent of brain injury is evident. However, we advocate caution when interpreting this result which may be artefactual due to a small number of deaths occurring after 30-days. More data over an extended number of years are required to test the robustness of this finding.

### Comparison with other studies

Several studies have demonstrated recent declines in stroke rates and mortality, although this experience is not universal.^30-39^ Two studies utilising administrative data from England^30^ and Sweden^31^ published in 2019, reported stroke cases from 2001 to 2010 and from 1994 to 2014, respectively. While neither study included subarachnoid haemorrhage nor reported statistics separately for intracerebral and ischaemic stroke, both reported reductions in stroke incidence and mortality. Our finding of unchanged risk of subarachnoid haemorrhage and declining mortality corroborates other findings.^36^ This analysis updates our earlier data linkage study reporting decreasing mortality for intracerebral^12^ and subarachnoid haemorrhage in NSW from 2001 to 2009,^13^ demonstrating sustained mortality reductions for these sub-types with more recent data.

Here and elsewhere,^30,32-35,37,38^ trends differed according to age when analysing individual age bands; however, we found no evidence that trends differed according to age when assessed using interaction terms. Analysis of individual age-bands is more susceptible to random variation, particularly in younger age-groups with relatively fewer cases, therefore such results should be interpreted cautiously.

### Strengths and limitations

Strengths of the study include whole-of-population continuous assessment of admissions and mortality over a nine-year period and data-linkage enabling individual level analyses. We cannot exclude the possibility of misclassification due to coding errors. Variability in coding over time may have occurred as the up-scaling of in-hospital stroke resources may have improved the identification and recording of stroke cases, although this effect would likely temper decreasing trends. As summarised previously,^8^ evidence demonstrates high levels of coding accuracy.

Unspecified strokes handled as ischaemic stroke improves the sensitivity of ischaemic stroke classification without affecting positive predictive value^15^ and we therefore followed recommendations to regard strokes coded as unspecified as ischaemic events. Here, 23% cases of all cases prior to applying exclusions were coded as unspecified strokes and we utilised more specific stroke codes, where recorded, to inform subtype classification. Any misclassification of ischaemic and intracerebral haemorrhage was minimised in analyses combining cases coded as such into a single category.

Fact of death registrations were extracted on the basis of date of registration and not date of death. Ascertainment of 30-day mortality would be complete as we accessed registered deaths at least one-year and up to nine years after admission. Our assessment of 365-day mortality may have been under-estimated if deaths occurring within one year for admissions in the last month of the study period had not been registered. In NSW, all deaths must be registered within seven days of burial or cremation and therefore any under-counting of 365-day mortality likely would be very slight.

We would not have captured deaths if these occurred in another state. However, there is no plausible reason to suggest out-of-state deaths have changed. Cases dying in emergency departments were captured. We did not include cases not presenting to hospital. Analyses are best seen as describing the burden of stroke on hospital services and outcomes in cases managed after reaching hospital. Reductions in stroke risk and improvements in outcomes may not generalise to all members of our community, specifically, First Nations people^35,40^ who experience an elevated risk of stroke.

## Conclusions

Improvements in stroke prevention may have reduced the risk of intracerebral haemorrhage. A lack of change in subarachnoid haemorrhage risk highlights ongoing difficulties for primary prevention. In contrast, stroke mortality after hospital admission has improved for all sub-types. This may be due to strokes becoming milder over time with more effective stroke risk prevention and/or successful translation of innovations in stroke management to reduce death and disability. Longer-term mortality in patients surviving the acute period is unchanged and may be increasing in subarachnoid haemorrhage, warranting investment in sub-acute management and post-discharge care and secondary prevention.

## Data Availability

Data utilised for these analyses are third party-owned. The terms of the agreements between researchers and data custodians preclude data sharing. Requests from qualified researchers to access third-party owned datasets used in these analyses may be sent to the Centre for Health Record Linkage.

## Conflicts of Interest

John Worthington serves as a Board Member, NSW Ministry of Health, Bureau of Health Information (unpaid position). The Bureau of Health Information utilises linked and unlinked routinely collected health data to evaluate patient outcomes in NSW hospitals, including stroke. JW was a paid consultant for the Agency for Clinical Innovation, NSW Ministry of Health (2014-2016) to lead a project to reduce unwarranted clinical variation in stroke across the NSW Health Service.

## Funding

New South Wales Ministry of Health, Office for Health and Medical Research, New South Wales Neurological Conditions Translational Research Grants Program.

## Acknowledgements

We gratefully acknowledge the Centre for Health Record Linkage for data linkage services and the NSW Ministry of Health for supplying data for these analyses. Data analysed as part of these analyses are third-party owned. Requests to access these data may be directed to the Centre for Health Record Linkage, NSW Ministry of Health.

## Key points

- In this system-wide analysis of 81,703 stroke admissions, rates of intracerebral haemorrhage and ischaemic stroke decreased from 2005 to 2013, while deaths within 30-days decreased for all sub-types.
- Rates of 365-day mortality amongst 30-day survivors were unchanged for ischaemic stroke and intracerebral haemorrhage yet increased for subarachnoid haemorrhage.
- Improved management of hypertension may have reduced intracerebral haemorrhage risk, while a trend towards milder strokes and wider implementation of evidence-based acute care may have underpinned improvements in survival.
- Public health initiatives should focus on both primary and secondary prevention to reduce stroke risk and improve longer-term survival.
- Policy initiatives to upscale and intensify evidence-based care for managing stroke is indicated to improve outcomes.

